# High risk injection drug use and uptake of HIV prevention and treatment services among people who inject drugs in KwaZulu-Natal, South Africa

**DOI:** 10.1101/2023.01.16.23284613

**Authors:** Brian C. Zanoni, Cecilia Milford, Kedibone Sithole, Nzwakie Mosery, Michael Wilson, Shannon Bosman, Jennifer Smit

## Abstract

The use of injection drugs in South Africa is increasing. HIV prevention, treatment and addiction services for people who inject drugs (PWID) in South Africa are not well documented. We conducted a mixed-methods study to understand current drug use practices and access to HIV prevention and treatment services for PWID in KwaZulu-Natal, South Africa. We used respondent-driven sampling to recruit 45 people who reported injecting opiates within the past 6 months from Durban, KwaZulu-Natal, South Africa. We found high rates of practices that increase HIV/viral hepatitis risk including the use of shared needles (43%) and direct blood injections (bluetoothing) (18%). Despite 35% of participants living with HIV, only 40% accessed antiretroviral therapy within the past year, and one accessed PrEP. None of the participants reported ever testing for Hepatitis C.

## Introduction

South Africa has an estimated 7.9 million (95% CI: 7.1 million – 8.8 million) individuals living with HIV, and KwaZulu-Natal has the highest HIV prevalence in the country. (1) In South Africa, there are approximately 5 million people accessing antiretroviral therapy (ART), making it the largest national antiretroviral program in the world. (1) However, hidden populations remain that are not reached in the HIV prevention and care continuum, and efforts to expand HIV prevention, HIV testing and linkage to care services in sub-Saharan Africa have largely targeted non-injection drug using communities. (2, 3) Before a recent increase in people who inject drugs (PWID) in South Africa, it was estimated that 21% of PWID in South Africa were living with HIV (2, 4) compared to 14% in the general population. (1) In the general population 74% of people living with HIV are accessing ART while only 40.5% of PWID and living with HIV are accessing ART.(2) A recent increase in injection drug use, coupled with poor access to addiction services in the forms of needle exchanges or medication assisted therapy (such as methadone or buprenorphine), has led to the increase in a sequestered population that has potential to reverse many of the investments and gains in public health access to HIV prevention and treatment services. (5) The population of PWID in South Africa spans groups of homeless, sex workers, and working-class individuals, bridging multiple social networks contributing to the potential increase in HIV incidence among individuals not targeted in typical HIV continuum interventions. (6)

Whoonga, an opiate-based drug of abuse, has been present in KwaZulu-Natal for more than 10 years, yet has only recently seen an increase in injection use. (7, 8) In the past, Whoonga was typically smoked; however, intravenous and subcutaneous administration has become more prevalent. (9) The recent increase in cases of infective endocarditis in South Africa is indicative of an increase in injection drug use. (5, 7, 8) In addition, the dangerous practice of “bluetoothing” in which blood is withdrawn from one individual who has recently injected a drug and directly injected intravenously into another person, has been reported. (10, 11) Called flash-blooding in other African countries, this practice is often performed in settings of poverty and poor access to needles or needle exchange programs. (12-14) This practice, in addition to needle sharing, has potential to increase the risk of HIV and viral hepatitis among PWID and spill over into the general population through sexual networks. (15-18)

Given the limited data on the population of individuals using injection drugs in KwaZulu-Natal, South Africa, we conducted a mixed-methods, respondent-driven study to understand drug use practices and current access to health care in preparation for targeted prevention and treatment interventions.

## Methods

We used respondent driven sampling (RDS) to recruit individuals aged 18 years or older, with self-described use of injection drugs within the last 6 months, and currently living in KwaZulu-Natal, South Africa. We excluded individuals who did not speak either English or isiZulu, who were severely or visibly intoxicated, or those with severe mental or physical illness preventing participation in informed consent procedures.

We recruited three initial seed individuals (who also reported injection drug use in the past 6 months) who were attending a harm reduction center in Durban, South Africa. These three seed individuals were encouraged to recruit up to three other PWID from their individual social network. Each additional participant was also asked to recruit up to three different individuals from their own social network, until we reached a total sample of 45 participants. After providing written informed consent, participants completed a facilitated questionnaire that collected information on sociodemographics, drug use, sexual behavior and network characteristics, HIV testing practices, and use of HIV prevention or treatment services. Interviewers entered data directly into a REDCap database as questions were answered. (19)

We assessed the frequency, type, and methods of drug use, information on needle procurement, use, and sharing using the National HIV Behavior Surveillance System; (20) and WHO ASSIST. (21) HIV risk assessment including sex work, number of sexual partners, condom use and frequency of condom use within the past 3 months was assessed by the Texas Christian University HIV/Hepatitis Risk Assessment. (22) We also evaluated access to health care by exploring knowledge of HIV status, HIV testing in the past 12 months, access and use of medical services in the past 12 months, knowledge and acceptability of HIV self-testing.

For this descriptive analysis we used standard summary statistics (e.g. counts/percentages; median and interquartile range of continuous measures). Basic descriptive data analyses were performed using REDCap. Results of additional in-depth interviews are reported separately.

This study was approved by the Institutional review Boards of Emory University, the University of Witwatersrand and the KwaZulu-Natal National Department of Health.

## Results

We interviewed 45 individuals in Durban, South Africa who reported recent injection drug use using respondent-driven sampling from November 1, 2021 to February 8, 2022. Participants had a median age of 28.5 years (IQR 26.6 – 32.3) and were predominantly male (58%; n=26), heterosexual (91%; n=41), had completed secondary education (87%; n=39), and were currently (76%; n=32) or recently (92%; n=42) homeless as indicated by **Table 1**. The median reported starting age of injecting drug use was 22 years (IQR 19 – 26). The majority of participants did not use alcohol (64%; n=29) but reported use of other drugs (53%; n=24). Most participants (93%; n=42) reported use of injection drugs within the last month, with 91% (n=41) reporting averaging more than one injection per day. All participants (100%) reported daily use of Whoonga. Opioid overdoses were personally experienced in 22% (n=10) of participants in the last year, with a median of 3.5 (IQR 2 – 4.75) individual episodes of overdosing. Sixty four percent (n=28) knew others who had experienced a lethal overdose in the last year.

**Table 1:** Descriptive statistics of individuals recruited through respondent-driven sampling and self-reporting recent injection drug use in KwaZulu-Natal, South Africa.

| Characteristic | Participants n (%)<br>N=45 |
| --- | --- |
| **Demographics** |  |
| Median Age (years)(IQR) | 28.5 (26.6 – 32.3) |
| Male | 26 (58%) |
| Heterosexual | 41 (91%) |
| Single | 17 (39%) |
| Long term partner but not married and not living together | 26 (59%) |
| Several casual partners | 2 (5%) |
| South African citizen | 45 (100%) |
| **Education** |  |
| Primary education | 4 (9%) |
| Secondary education | 39 (87%) |
| Tertiary level | 2 (4%) |
| Median grade completed (IQR) | 10 (10 – 11) |
| **Employment** |  |
| Full time employed | 1 (2%) |
| Employed part time | 11 (24%) |
| Unemployed | 17 (38%) |
| **Housing** |  |
| Homeless in the past 12 months | 42 (93%) |
| Currently homeless | 32 (76%) |
| **Alcohol Use** |  |
| No alcohol in the past year | 29 (64%) |
| Binge drinking more than once a month | 10 (22%) |
| **Injection drug use** |  |
| Used injection drugs | 45 (100%) |
| Median age at first use (years) (IQR) | 22 (19 – 26) |
| Last used injection drugs in the past month | 42 (93%) |
| Injecting more than once a day | 41 (91%) |
| Injecting daily | 2 (4%) |
| Injecting more than once a week | 2 (4%) |
| Using Whoonga/heroin daily or more | 45 (100%) |
| Had an opioid overdose (in last year) | 10 (22%) |
| Median number of opioid overdoses (IQR) (in last year) | 3.5 (2 – 4.75) |
| Know of others who experienced lethal opioid overdose (in last year) | 29 (64%) |
| Median overdose deaths known about (in last year) | 2 (1 -3) |
| Know of others with non-lethal opioid overdose (in last year) | 28 (62%) |
| **Most common place acquired needles** |  |
| Needle exchange or NGO | 29 (64%) |
| Friend / acquaintance | 4 (11%) |
| Pharmacy | 1 (4%) |
| **Dispose of needle** |  |
| Needle exchange or NGO | 25 (56%) |
| On the street | 8 (8%) |
| Trash | 7 (16%) |
| **High risk Injection Drug Use** |  |
| New / unused Needle use in last 12 months |  |
| Always | 14 (31%) |
| Sometimes | 22 (49%) |
| Rarely | 9 (20%) |
| Never | 0 (0%) |
| Median number of people shared needles with in last 12 months (IQR) | 0 (0 -3) |
| Median number of people shared other drug-use material with in last 12 months (IQR) | 3 (0 – 5) |
| Median number of people shared drugs with in the last 12 months (IQR) | 3 (1 – 4) |
| Used a shared needle in the past 12 months | 19 (42%) |
| Used shared material in last 12 months | 33 (73%) |
| Bluetoothing in the last 12 months | 8 (18%) |
| Re-used needle from someone else at last injection | 10 (22%) |
| Bluetoothing at last injection | 2 (4%) |
| Did not dispose of needle at last injection | 29 (64%) |
| Injected with known HIV+ individual | 6 (13%) |
| Other drug use in last year | 24 (53%) |
| **Healthcare Services** |  |
| Participated in a drug treatment program | 27 (60%) |
| Ever tested for HIV | 43 (96%) |
| Known to be living with HIV | 15 (35%) |
| Accessing ART in the last 12 months | 6 (40%) |
| Median number of HIV tests in last 2 years | 3 (1 – 8.5) |
| Never tested for Hepatitis C | 45 (100%) |
| Currently taking PrEP | 1 (2%) |
| Interested in PrEP | 24 (53%) |

All participants (100%) reported ever re-using needles or equipment with 42% (n=19) reporting use of shared needles in the past year and 73% (n=33) reported sharing other drug preparation or use materials. The majority accessed (64%; n=29) and disposed (56%; n=25) of their needles through needle exchange programs. Bluetoothing was practiced in 18% (n=8) of individuals in the last 12 months.

More than half (60%; n=27) had ever participated in a drug treatment program. The majority (96%; n=43) had ever tested for HIV. Thirty-five percent of individuals (n=15) were known to be living with HIV but only six (40%) reported accessing antiretroviral therapy within the past 12 months. Only one individual (2%) was taking pre-exposure prophylaxis (PrEP). None of the participants had ever tested for Hepatitis C.

## Discussion

In this study we found high rates of unsafe injection drug practices that included sharing and reusing needles and drug preparation materials, as well as the practice of directly injecting blood from an individual who had recently injected (bluetoothing). These practices, if widespread amongst PWID, coupled with the low uptake of treatment and preventative services described in this study, including HIV testing, PrEP and ART services, could negate some of the efforts made in the HIV prevention and treatment continuum of care in South Africa.

The current and substantial efforts to combat the HIV epidemic in South Africa do not adequately address the needs of PWID. Although efforts are increasing to routinely screen for alcohol use in HIV care across South Africa, (23) screening for drug use (and injection drug use specifically) is not routinely conducted, and treatment for drug use disorder is rarely integrated with HIV care services.

While the COVID-19 pandemic slowed progress globally to provide access to harm reduction and health services to people who use drugs, parts of South Africa saw a positive shift from a once, a prohibitionist stance to dealing with drug use disorder to one that supports the provision of evidence-based harm reduction practices, especially to low-income and homeless people who use drugs. Nationally, needle exchange programs, harm reduction services, and medication assisted treatment are often difficult to access either due to their high-priced, fee-for-service models or limited availability. (24) KwaZulu-Natal’s only needle and syringe exchange program in Durban was reinstated during the COVID-19 lockdown, and the country’s first low-threshold harm reduction center was started in Durban to provide daily opiate substitution therapy to more than 200 PWID. The limited resources available for harm reduction services have historically resulted in punishment of substance use as the predominant response by law enforcement to opiate addiction. (25) However after the COVID-19 lockdowns, Durban also saw a dramatic shift in the way that law enforcement engaged with people who use drugs, shifting the narrative of police as punitive to one of protectors and advocates of health services for PWID. (26, 27) Continued work is needed to reduce ongoing stigma towards PWID which limits access to HIV prevention services and harm reduction programs. (28, 29)

Without the availability of widespread interventions, injection drug use has continued to increase in South Africa with a prevalence of 0.6% in a population based survey in 2012 with 0.2% reporting Whoonga use. (30) However, small pilot studies have shown high retention rates, opiate reduction, and improvement in mental health in individuals accessing methadone treatment in South Africa. (31) Nevertheless, the lack of routine hepatitis C testing and access to PrEP indicate significant gaps in service delivery. Integration of harm reduction services into routine healthcare could impact the rising use of opiates and the HIV epidemic in South Africa. The major limitations of this study include the small sample size and descriptive nature of this preliminary study. In addition, this study was conducted during the COVID-19 pandemic when services were interrupted by various lockdown policies. During this time, there was also an influx of new people into the city who may not have been familiar with available services. Unsafe drug use practices may have occurred as a result of disruption of needles and other safe use supplies during lockdown. However, we are encouraged by the ability to recruit a difficult to reach population through respondent-driven sampling over a short period of time (3 months) and using only 3 initial seed individuals.

## Conclusion

The increased prevalence of injection drug use in South Africa along with unsafe injection practices and low uptake of preventative and treatment services, in particular HIV related services, has potential to reverse the significant gains made in HIV prevention services. Integration of addition services, medication assisted treatment and access to HIV prevention and treatment services could improve the lives of PWID and reduce the burden of HIV in this population.

## Data Availability

All relevant data are within the manuscript and its Supporting Information files.

## Author’s Contributions

Dr. Zanoni conceptualized and designed the study, performed the data analysis, drafted the initial manuscript, reviewed and revised the manuscript and approved the final manuscript as submitted.

Kedibone Sithole performed the data collection, contributed to the drafting of the manuscript, critically reviewed the manuscript and approved the final manuscript as submitted.

Nzwakie Mosery performed the data collection, contributed to the drafting of the manuscript, critically reviewed the manuscript and approved the final manuscript as submitted.

Michael Wilson assisted with the conceptualization and design of the study, contributed to the data collection, and critically reviewed the manuscript, and approved the final manuscript as submitted.

Shannon Bosman contributed to the data collection, and critically reviewed the manuscript, and approved the final manuscript as submitted.

Cecilia Milford assisted with the conceptualization and design of the study, contributed to the analysis plan, reviewed and revised the manuscript, and approved the final manuscript as submitted.

Jennifer Smit assisted with the conceptualization and design of the study, contributed to the analysis plan, reviewed and revised the manuscript, and approved the final manuscript as submitted.

